# Predicted behavioural responses to tobacco product regulations: findings from an online survey of Australian adults who smoke

**DOI:** 10.1101/2025.06.09.25329296

**Authors:** Claudia Gascoyne, Tegan Nuss, Ashleigh Haynes, Melanie Wakefield, Sarah Durkin, Emily Brennan

## Abstract

**Background:** Regulating features of tobacco products can reduce the appeal and addictiveness of smoking. We examined predicted behavioural responses to bans on i) filter ventilation (FV), ii) regular nicotine content (RNC), and iii) menthol and flavour crushballs (MFC), as well as perceived rationales for these bans.

**Methods:** Australian adults who smoked at least weekly were recruited from an online research panel to participate in a cross-sectional survey and were allocated to either the FV (n=301), RNC (n=335), or MFC (n=298) study arm, depending on current product use. Respondents viewed a basic description of each ban before completing measures of predicted behavioural responses and perceived rationale.

**Results:** The proportion of respondents who reported being likely to try to quit smoking in response to the product regulations was 25.9% in the FV arm, 18.5% in the RNC arm, and 13.4% in the MFC arm. However, in each study arm, respondents more commonly predicted that they would switch to tobacco products that would remain available after the ban (FV: 33.6%; RNC: 37.0%; MFC: 51.3%), and some reported being likely to switch to e-cigarettes (FV: 14.6%; RNC: 11.6%; MFC: 23.2%). The majority of respondents reported either misperceptions or uncertainty about the rationale for the bans (FV: 69.8%; RNC: 81.5%; MFC: 62.4%).

**Conclusion:** A higher proportion of people who smoke anticipate that they would respond to tobacco product bans by switching to a different product, rather than using the ban as an opportunity to quit. Future studies should experimentally investigate whether public health messaging about these product regulation measures can be used to encourage greater rates of quitting by minimising switching to available products, including e-cigarettes, and to minimise misperceptions by increasing awareness of the rationale for each product regulation measure.

## INTRODUCTION

Smoking remains the leading cause of preventable death in Australia,[1] and there are large disparities in smoking prevalence across social, geographical, and cultural groups, as well as between those with and without comorbidities.[2–6] To reduce the appeal and addictiveness of smoking, the Australian Government’s National Tobacco Strategy 2023-2030 proposes several actions to regulate the contents, design and appearance of tobacco products in line with Article 9 of the WHO Framework Convention on Tobacco Control.[7, 8] The *Public Health (Tobacco and Other Products) Act 2024*, and the Regulations that accompany the Act, includes a ban on the use of certain flavouring additives including menthol, and standardisation of the design and appearance of cigarette filters, including by banning filters with flavour crushballs. Each of these policies is to be in full effect by July 1 2025.[9] Other product regulations recommended in Article 9 include a ban on filter ventilation (which refers to perforations in cigarette tipping paper which mixes air with the smoke, reducing its harshness) and limiting the nicotine content in all available tobacco products to levels lower than typically present.

The regulation of certain tobacco product characteristics has proven to be an effective strategy to promote quit attempts among consumers of the regulated product. Following the ban on menthol tobacco products in Canada in 2017, longitudinal data showed that those who smoked menthol products reported higher rates of quit attempts and quit success compared to those who smoked non-menthol products.[10, 11] Similarly, longitudinal data from The Netherlands showed a decrease in the use of menthol products and an increase in quit attempts among those who consumed these products following the European Union menthol ban in 2020.[12] Despite these promising data, there is also evidence that some people who smoke may respond to new restrictions by switching to a tobacco product that remains available on the market. For example, one year after the ban in Canada, 22% of those who smoked menthol cigarettes had quit altogether, while 59% had switched to non-menthol cigarettes.[10]

One factor that may contribute to higher rates of product switching rather than quitting is misperceptions about the relative harmfulness of banned and available products. For instance, studies have shown that when informed about potential constraints on the amount of nicotine allowed in tobacco products, some people who smoke incorrectly believed that the new low nicotine content cigarettes (i.e., the available product) would be less harmful than regular nicotine content cigarettes (i.e., the banned product)[13–15], and this misperception was associated with a reduced likelihood of quitting in response to the regulation of nicotine content.[15] It will therefore be essential to establish strategies to minimise consumer misperceptions around the rationale for tobacco product regulations in order to maximise the potential for these regulations to encourage smoking cessation. However, the prevalence of misperceptions related to various tobacco product regulations among Australian adults who smoke is not yet known.

As governments move towards regulating selected tobacco product attributes, there is a need to identify opportunities to minimise potential unintended consequences in order to maximise the public health impact of these regulations. This study aimed to examine predicted behavioural responses to three different tobacco product regulation measures among adults who smoke. We also examined perceived rationales for why these product regulation measures would be introduced to assess the prevalence of misperceptions about these measures.

## METHODS

### Survey sample and procedure

An online cross-sectional survey was conducted between December 14 and 18, 2022. The survey had three study arms, with each respondent assigned to one of the arms: i) filter ventilation (FV), featuring a hypothetical ban on filter ventilated tobacco products; ii) regular nicotine content (RNC), featuring a hypothetical limit on the amount of nicotine allowed in tobacco products; and iii) menthol/flavoured crushball (MFC), featuring the ban on the use of flavour and menthol in tobacco products being implemented in Australia from mid-2024 (with menthol and flavoured crushball products most likely to begin disappearing from the market in early 2025).

All respondents were Australian residents aged between 18 and 69 years recruited from an opt-in non-probability online research panel maintained by a data collection agency accredited by the International Organization for Standardization (ISO). After completing socio-demographic and smoking behaviour measures to assess their eligibility for the study and for each study arm, respondents were allocated to a study arm based on their current smoking behaviours. Respondents were eligible for the FV study arm if they currently smoked filter ventilated tailor-made cigarettes (defined for purposes of the study as being any colour variant, excluding red and black variants) at least weekly. Respondents were eligible for the RNC study arm if they consumed any type of tobacco product at least weekly, including filter ventilated tailor-made cigarettes and MFC cigarettes. Respondents were eligible for the MFC study arm if they currently smoked menthol and/or flavoured crushball tailor-made or roll-your-own cigarettes at least weekly. Those who were eligible for more than one study arm were allocated based on ‘least-fill’; that is, they were allocated to the arm that had the lowest number of completions to ensure an even representation of respondents in each arm. Soft quotas were applied to ensure that each study arm was approximately representative in terms of gender, age group, and product use (FV study arm: 50%/50% male/female, 50%/50% 18-39/40-69 years; RNC study arm: 50%/50% male/female, 50%/50% 18-39/40-69 years, 25% weekly consumer of MFC cigarettes, 50% weekly consumer of FV tailor-made cigarettes, 25% weekly consumer of any other tobacco product; MFC study arm: 50%/50% male/female, 60%/40% 18-39/40-69 years). Respondents were ineligible to participate if they reported having completed an online questionnaire about cigarette smoking or other tobacco products more than ten times in the past six months.

Within each study arm, respondents were asked to read a brief description of the tobacco product regulation measure relevant to that study arm and were told that this potential change in the law would be described in the form of a short news article (see Table 1). To ensure that respondents were aware that the tobacco product regulations featured in the FV and RNC study arms were hypothetical, respondents in these two study arms were also informed that governments in some other countries like New Zealand and the United States are considering making changes to the law about which tobacco products can be sold, and that we were interested in what they think they would do if this law was also passed in Australia. As part of their debriefing at the end of the study, these respondents were also reminded that the scenario described was hypothetical.

**Table 1.** Descriptions of the tobacco product regulations shown to the study sample, by study arm.

| Study arm | Description |
| --- | --- |
| Filter ventilation | **Australian Government to ban filter ventilated cigarettes**<br>In three months' time, tailor-made cigarettes sold in Australia will no longer be able to use **filter ventilation**. Filter ventilation refers to tiny holes being placed around the filter. Air is drawn in through these tiny holes, diluting the smoke and making it feel lighter and smoother. This means that the only tailor-made cigarettes available will be very strong and harsh, like those currently labelled "red". |
| Regular nicotine content | **Australian Government to restrict the amount of nicotine in tobacco products**<br>In three months' time, cigarettes and roll-your-own tobacco that contain regular amounts of nicotine will no longer be allowed to be sold in Australia. This means that the only tobacco products available will be new products in which the amount of nicotine is dramatically reduced so that the products are no longer addictive. These tobacco products are called "Very Low Nicotine Cigarettes" or "Very Low Nicotine Tobacco". |
| Menthol/flavoured crushball | The Australian government recently announced plans to change the law so that all tobacco products that contain **menthol in the tobacco** will no longer be allowed to be sold in Australia. They are also ending the sale of cigarettes that contain **flavoured crushball capsules in the filter**, and **filters with flavoured crushball capsules** that are used for roll-your-own cigarettes. It is likely that this new law will come into force in 2024. |

### Measures

Following exposure to the product regulation announcement, two primary outcomes were measured: i) predicted behavioural response to the announcement, and ii) perceived rationale for the regulatory change.

Predicted behavioural responses were measured by asking respondents what they would be most likely to do if the regulatory changes were introduced in Australia. Respondents were asked to select one of 11 response options provided: smoke tobacco products that would still be available for sale, use various types of e-cigarettes, find a way to get the banned product even though it would no longer be legally available, modify available products or the way they are consumed to make it seem more like the banned product, try to quit smoking, or don’t know / can’t say. Respondents were also able to select ‘other’ and specify in their own words their most likely response.

To assess their perceived rationale for the regulatory change, respondents were asked why they think the government would introduce this measure. In each study arm, respondents were provided with seven response options, including at least one correct response (e.g., ‘nicotine is the substance that makes tobacco products addictive’), various incorrect responses representing misperceptions (e.g., ‘nicotine causes cancer’; see Table 4 for full list of response options), and a ‘don’t know / can’t say’ option.

Given that the Australian Government announced their plans to ban menthol and flavoured crushballs on November 30, 2022,[17] a few weeks prior to data collection, respondents in the MFC study arm were additionally asked whether they had previously heard about these plans (‘yes’, ‘no’, ‘don’t know / can’t say’).

### Statistical analysis

Data were analysed using Stata MP version 16.1. Within each study arm, descriptive statistics are reported overall for the two primary outcome measures: i) predicted behavioural response to the product regulation measure, and ii) perceived rationale for the product regulation measure. Multivariate logistic regression models were used within the MFC study arm with prior awareness of the Australian Government’s plan to ban menthol and flavoured crushballs included as a covariate (prior awareness cf. no prior awareness or don’t know / can’t say). A significance level of *p*<0.05 was accepted throughout. All proportions reported within the FV and RNC study arms are unadjusted, while all proportions reported within the MFC study arm are adjusted for prior awareness of the menthol and flavour crushball ban.

## RESULTS

### Participants

In total, 301 respondents were allocated to the FV arm, 335 were allocated to the RNC arm, and 298 were allocated to the MFC arm. The socio-demographic characteristics of these samples are shown in Table 2.

**Table 2.** Study sample characteristics by study arm.

|  | Filter ventilation,<br>N (%) | Regular nicotine<br>content, N (%) | Menthol / flavour<br>crushball, N (%) |
| --- | --- | --- | --- |
| *Total* | 301 (100.0) | 335 (100.0) | 298 (100.0) |
| *Age group* |  |  |  |
| 18-39 years | 144 (47.8) | 158 (47.2) | 179 (60.1) |
| 40-69 years | 157 (52.2) | 177 (52.8) | 119 (39.9) |
| *Gender* |  |  |  |
| Man or male | 146 (48.5) | 160 (47.8) | 141 (47.3) |
| Woman or female | 155 (51.5) | 174 (51.9) | 156 (52.3) |
| Other term or prefer not to say | 0 (0.0) | 1 (0.3) | 1 (0.3) |
| *Socio-economic area* |  |  |  |
| Low (1-40%) | 128 (42.5) | 146 (43.7) | 125 (41.9) |
| Mid (41-80%) | 124 (41.2) | 137 (41.0) | 121 (40.6) |
| High (81-100%) | 49 (16.3) | 51 (15.3) | 52 (17.4) |
| *Geographic location* |  |  |  |
| Metropolitan | 220 (73.1) | 220 (65.7) | 221 (74.2) |
| Regional | 81 (26.9) | 115 (34.3) | 77 (25.8) |
| *Identifies as Aboriginal or Torres Strait Islander* |  |  |  |
| Yes | 11 (3.7) | 21 (6.3) | 35 (11.7) |
| No | 290 (96.3) | 314 (93.7) | 263 (88.3) |
| *Highest level of education* |  |  |  |
| Not completed tertiary education | 165 (55.0) | 219 (65.8) | 137 (46.1) |
| Completed tertiary education | 135 (45.0) | 114 (34.2) | 160 (53.9) |
| *Health care or pensioner concession card holder* |  |  |  |
| Yes | 104 (34.6) | 133 (39.7) | 109 (36.6) |
| No | 197 (65.4) | 202 (60.3) | 189 (63.4) |
| *Number of 24+ hour quit attempts in past month* |  |  |  |
| None or don't know/can't say | 204 (67.8) | 235 (70.1) | 142 (47.7) |
| At least once | 97 (32.2) | 100 (29.9) | 156 (52.3) |
| *Quit intentions* |  |  |  |
| Not considering quitting in next 6 months | 102 (33.9) | 135 (40.3) | 75 (25.2) |
| Considering quitting in next 6 months | 150 (49.8) | 156 (46.6) | 170 (57.0) |
| Plans to quit in next month | 49 (16.3) | 44 (13.1) | 53 (17.8) |
| *Frequency of TM cigarette consumption* |  |  |  |
| Less than weekly or not at all | 0 (0.0) | 52 (15.6) | 13 (4.4) |
| Weekly | 301 (100.0) | 282 (84.4) | 285 (95.6) |
| *Frequency of RYO cigarette consumption* |  |  |  |
| Less than weekly or not at all | 196 (66.0) | 170 (51.2) | 117 (39.5) |
| Weekly | 101 (34.0) | 162 (48.8) | 179 (60.5) |

Notes: Proportions are rounded so may not sum to 100.0%. Socio-economic area could not be computed for n=1 in the regular nicotine content arm. No response was provided for highest level of education for n=1 in the filter ventilation arm, n=2 in the regular nicotine content arm, and n=1 in the menthol / flavour crushball arm. Frequency of TM cigarette consumption was not reported by n=1 in the regular nicotine content arm. Frequency of RYO cigarette consumption was not reported by n=4 in the filter ventilation arm, n=3 in the regular nicotine content arm, and n=2 in the menthol / flavour crushball arm. Abbreviations: TM=tailor-made; RYO=roll-your-own.

### Filter Ventilation arm

Table 3 shows that 25.9% of this sample reported they would most likely try to quit smoking in response to a ban on filter-ventilated tobacco products. Approximately one in three (33.6%) reported they would smoke available tobacco products, including roll-your-own cigarettes (18.6%) and unventilated tailor-made cigarettes (15.0%), and 14.6% reported they would use either flavoured or unflavoured e-cigarettes. Around one in eight (13.3%) reported being unsure what they would do in response to the regulation.

**Table 3.** Self-reported predicted behavioural response to product regulations, by study arm.

|  | Overall, % |
| --- | --- |
| *Filter ventilation arm* |  |
| Try to quit smoking | 25.9 |
| Smoke unventilated tailor-made cigarettes | 15.0 |
| Smoke roll-your-own cigarettes | 18.6 |
| Use flavoured vapes / e-cigarettes | 13.3 |
| Use unflavoured vapes / e-cigarettes | 1.3 |
| Modify unventilated tailor-made cigarettes | 5.3 |
| Change the way unventilated tailor-made cigarettes are smoked | 3.3 |
| Find a way to get filter ventilated cigarettes | 4.0 |
| Don't know / Can't say | 13.3 |
| *Regular nicotine content arm* |  |
| Try to quit smoking | 18.5 |
| Smoke Very Low Nicotine tailor-made cigarettes | 23.6 |
| Smoke Very Low Nicotine roll-your-own cigarettes | 13.4 |
| Use flavoured vapes / e-cigarettes | 10.5 |
| Use unflavoured vapes / e-cigarettes | 1.2 |
| Modify the Very Low Nicotine cigarettes/tobacco to make them more like the cigarettes/tobacco with regular amounts of nicotine | 3.0 |
| Change the way you smoke Very Low Nicotine cigarettes/tobacco to make it feel like you're smoking cigarettes/tobacco with nicotine | 4.2 |
| Find a way to get tobacco products with regular amounts of nicotine, even if they are no longer legally for sale | 16.4 |
| Don't know / Can't say | 9.3 |
| *Menthol /flavour crushball arm* |  |
| Try to quit smoking | 13.4 |
| Smoke non-menthol tailor-made cigarettes with regular filters | 33.6 |
| Smoke non-menthol roll-your-own cigarettes with regular filters | 17.8 |
| Use menthol or mint flavoured vapes / e-cigarettes | 9.1 |
| Use other flavoured vapes / e-cigarettes | 8.1 |
| Use unflavoured vapes / e-cigarettes | 6.0 |
| Modify the cigarettes that are available to make them taste more like my preferred cigarettes | 2.3 |
| Find a way to get tobacco products that contain menthol, or flavoured crushball cigarettes, even if they are no longer legally for sale | 5.7 |
| Don't know / Can't say | 4.0 |

Of this sample, 30.2% held only accurate perceptions about the FV ban rationale, while 69.8% held at least one misperception or were unsure about the reasons for the ban. Specifically, 38.5% correctly thought that the government would implement such regulations because the lighter and smoother smoke created by filter ventilated tailor-made cigarettes misleads smokers into believing they are less harmful than unventilated tailor-made cigarettes, even though they are just as harmful (see Table 4). In addition, 24.3% correctly thought that it was because the tobacco industry has promoted filter ventilated cigarettes as being less harmful than unventilated tailor-made cigarettes. The most frequently selected incorrect response was that the government earns more tax on unventilated cigarettes (24.6%). Almost one-quarter incorrectly thought that filter ventilated cigarettes were being banned because they are more harmful than either unventilated cigarettes (9.0%) or all other tobacco products (13.3%). Just over one-quarter (28.2%) were unsure about the reason for the ban.

**Table 4.** Self-reported perceived rationale for product regulations, by study arm.

|  | Overall, % |
| --- | --- |
| *Filter ventilation arm* |  |
| Accurate rationale |  |
| The lighter and smoother smoke created by filter ventilated tailor-made cigarettes misleads smokers into believing they are less harmful than unventilated tailor-made cigarettes | 38.5 |
| The tobacco industry has promoted filter ventilated cigarettes as being less harmful than unventilated tailor-made cigarettes | 24.3 |
| Inaccurate rationale |  |
| The government earns more tax on unventilated cigarettes | 24.6 |
| Filter ventilated tailor-made cigarettes are more harmful than all other tobacco products | 13.3 |
| Filter ventilated cigarettes are more damaging to the environment than unventilated cigarettes | 12.0 |
| Filter ventilated tailor-made cigarettes are more harmful than unventilated tailor-made cigarettes | 9.0 |
| Don't know / Can't say | 28.2 |
| *Regular nicotine content arm* |  |
| Accurate rationale |  |
| Nicotine is the substance that makes tobacco products addictive | 49.9 |
| Inaccurate rationale |  |
| The nicotine in tobacco is responsible for most diseases caused by smoking | 35.5 |
| Nicotine causes cancer | 32.8 |
| The government can earn more tax on Very Low Nicotine cigarettes/tobacco | 29.0 |
| Nicotine causes heart disease | 28.7 |
| Cigarettes/tobacco with regular levels of nicotine are more damaging to the environment than Very Low Nicotine cigarettes/tobacco | 18.5 |
| Don't know / Can't say | 14.3 |
| *Menthol / flavour crushball arm* |  |
| Accurate rationale |  |
| The fresher/smooth smoke created by menthol and crushball cigarettes misleads smokers into believing they are less harmful than other tobacco products | 42.3 |
| The tobacco industry has promoted menthol and crushball cigarettes as being less harmful than other tobacco products | 29.2 |
| Menthol and crushball cigarettes are more addictive than other tobacco products | 23.8 |
| Inaccurate rationale |  |
| Menthol and crushball cigarettes are more harmful than other tobacco products | 21.1 |
| Menthol and crushball cigarettes are more damaging to the environment than other tobacco products | 18.5 |
| The government earns more tax on non-menthol cigarettes and cigarettes that do not contain crushballs | 18.5 |
| Don't know / Can't say | 19.1 |

### Regular Nicotine Content arm

As shown in Table 3, 18.5% of this sample reported they would most likely try to quit smoking in response to a limit on the amount of nicotine allowed in tobacco products. More than one-third (37.0%) reported they would smoke available tobacco products, including Very Low Nicotine roll-your-own (13.4%) or tailor-made cigarettes (23.6%), and 11.6% reported they would use either flavoured or unflavoured e-cigarettes. Approximately one in ten (9.3%) reported being unsure what they would do in response to this regulation.

Of this sample, 18.5% held only accurate perceptions about the rationale for limiting the amount of nicotine allowed in tobacco products, while 81.5% held at least one misperception or were unsure. Almost half (49.9%) correctly thought that the government would implement such regulations because nicotine is the substance that makes tobacco products addictive (see Table 4), and this was the most common response. Approximately one in three incorrectly thought the reason was that the nicotine in tobacco is responsible for most diseases caused by smoking (35.5%), nicotine causes cancer (32.8%), or nicotine causes heart disease (28.7%), while 14.3% were unsure about the reason for the ban.

### Menthol and Flavour Crushball arm

After adjusting for prior awareness of the MFC ban (reported by 33.2% of the sample), 13.4% reported they would most likely try to quit smoking in response to a ban on menthol and flavour crushball tobacco products (Table 3). More than half (51.3%) reported they would smoke available tobacco products, including non-menthol tailor-made (33.6%) or roll-your-own (17.8%) cigarettes with regular filters, and almost a quarter (23.2%) reported they would use either menthol flavoured, other flavoured, or unflavoured e-cigarettes. A minority (4.0%) reported being unsure what they would do in response to the regulation.

Of this sample, 37.6% held only accurate perceptions about the MFC ban rationale, while 62.4% held at least one misperception or were unsure. As shown in Table 4, approximately two in five respondents (42.3%) correctly thought that the government would implement this regulation because the fresher or smoother smoke created by menthol and crushball cigarettes misleads smokers into believing they are less harmful than other tobacco products, even though they are just as harmful, and this was the most common response. Slightly fewer correctly thought that the rationale for the MFC ban was that the tobacco industry has promoted menthol and crushball cigarettes as being less harmful than other tobacco products, even though they are just as harmful (29.2%), and that menthol and crushball cigarettes are more addictive than other tobacco products (23.8%). Just over one-fifth incorrectly attributed the ban to menthol and crushball cigarettes being more harmful than other tobacco products (21.1%), while 19.1% were unsure why the government would implement these regulatory changes.

## DISCUSSION

Overall, the study findings indicate that fewer than one-third of people who smoke anticipate that they would quit in response to three potential product regulation measures intended to reduce the appeal and/or addictiveness of tobacco products. Rather, in each study arm, respondents more commonly predicted that they would use the main available tobacco products rather than try to quit, and at least one in ten expected that they would use e-cigarettes following implementation of the ban. Our study also identified that the majority of respondents within each study arm reported either misperceptions or uncertainty about the rationale for the ban. Taken together, these findings indicate a substantial potential to encourage greater rates of quitting in response to three product regulation measures with a need for clear public messaging to educate people who smoke about the reasons for these product regulation measures.

Our findings suggest that a ban on filter ventilated cigarettes has the potential to prompt quit attempts among over one-quarter of those who consume these products. However, given the substantial proportion of respondents (48.2%) who indicated they would use roll-your-own or unventilated tailor-made cigarettes or e-cigarettes (rather than quit smoking), it will be critical that a ban on filter ventilation be accompanied by messaging and other tobacco control interventions designed to reduce the appeal of available tobacco products and e-cigarettes. There was also a high prevalence of uncertainty regarding the government’s rationale for a hypothetical filter ventilation ban (28.2%), suggesting a need for public education that can improve understanding about what filter-ventilated cigarettes are and how they mislead about harms.[19–21]

Similarly, while almost one in five respondents in the RNC study arm indicated they would likely attempt to quit following a ban on regular nicotine content cigarettes, nearly half indicated they would smoke remaining tobacco products, specifically Very Low Nicotine roll-your-own or tailor-made cigarettes (37.0%) or e-cigarettes (11.7%). Inaccurate perceptions about the rationale for a regular nicotine content ban, including misunderstanding about the role of nicotine in tobacco products, may contribute to intentions to use Very Low Nicotine products rather than try to quit. While half of respondents in this study arm correctly identified the rationale for the ban (i.e., that nicotine is the substance that makes tobacco products addictive), it is concerning that approximately one in three selected each of the three incorrect response options about nicotine causing heart disease and cancer and being responsible for most of the diseases caused by smoking. This finding is in line with a 2017 systematic review and recent Australian qualitative research that found nicotine is commonly misperceived to cause cancer or be the primary source of harm in tobacco products.[22, 23] Hence, there is substantial potential to educate people who smoke that the other chemicals in tobacco smoke are responsible for most of the harms caused by smoking, not nicotine, which may help reduce intentions to switch to new Very Low Nicotine tobacco products and increase quit intentions. Notably, 16.4% of respondents in this study arm anticipated they would try to find a way to get tobacco products with regular amounts of nicotine, even if they are no longer legally for sale. This suggests an expectation that illicit tobacco products that are currently accessed in Australia by a small proportion of people who smoke would continue to be available.[24] In January 2024 the Australian Government announced plans for a multi-pronged strategy to be implemented over four years, which if successful, could substantially decrease the availability of illicit tobacco.[25]

Overall, 13.4% of respondents in the MFC study arm reported they would try to quit smoking in response to a ban on MFC cigarettes. This estimate is lower than provided by a recent review of the impact of menthol cigarette bans, which identified that 24% of people who smoked menthol tobacco products had quit smoking 1-2 years following a menthol ban and that this proportion was similar to hypothesized quitting prior to the ban (33%).[26] However, estimates of predicted quitting in response to a menthol ban varies greatly across studies, as identified in two separate reviews[26, 27], suggesting that the predicted quit rate in our study may too be highly variable. In particular, effects may be dependent on messaging that relevant audiences are exposed to prior to implementation of the MFC ban, particularly if these audiences have low existing awareness of the rationale for the ban. In light of this, it is worth noting that data for the current study were collected only a few weeks after the Australian Government announced plans for an MFC ban, so it is possible that the messaging included within and surrounding this announcement may have influenced predicted behavioural responses reported in our study.[17] The Australian public had not previously been exposed to any mass communication about the specific harms of menthol products, and the government’s announcement about the ban did not include detailed information about the rationale for the MFC ban nor an explicit call-to-action to quit smoking. Furthermore, the announcement also included details about a breadth of other tobacco control reforms to be introduced at the same time as the MFC ban. These factors may have contributed to confusion or even resistance to an MFC ban, although our estimate adjusted for prior awareness of the ban to limit the possibility of this bias. Furthermore, in contrast to the FV and RNC study arms, the proportion who reported they would try to quit in the MFC study arm may have been comparatively lower because less commitment is required to predict quitting in response to a hypothetical ban relative to a real ban. These various considerations suggest that our study findings may underestimate the actual proportion of people who would quit in response to an MFC ban, and therefore future studies should assess whether predicted behavioural responses vary depending on messaging strategy, with consideration of prior awareness of the MFC ban.

In response to the ban on MFC cigarettes, over half of MFC study arm respondents reported that they would use non-menthol cigarettes with regular filters, and almost one-quarter indicated that they would be most likely to use e-cigarettes. These findings loosely align with the recent review, which reported that up to 46% of people who smoke menthol cigarettes predicted they would switch to other tobacco products and 15-30% predicted they would switch to e-cigarettes.[27] There was evidence that misperceptions about the relative harmfulness of menthol and non-menthol cigarettes may be prevalent, with one in five respondents in this study arm incorrectly believing that the government plans to ban MFC cigarettes because these products are more harmful than other tobacco products. Taken together, these findings suggest that there are substantial opportunities to use public communication messages to prevent or correct misperceptions about the relative harm of menthol and non-menthol tobacco products, and to potentially influence behavioural responses as MFC bans are implemented. In particular, these findings suggest that messaging should aim to build upon existing levels of understanding that menthol and flavour crushball products are being banned because the fresher and smoother smoke misleads people who smoke into believing that these products are less harmful than they really are. In addition, it will be crucial to reinforce that tobacco products that remain available are just as harmful as those being banned and that e-cigarettes are not risk-free.

A number of study limitations should be noted. Firstly, this study did not aim to investigate which of the three assessed tobacco product regulations would be most effective at promoting quitting among people who smoke. Hence no recommendations are provided to this end, particularly given that the level of detail provided to respondents about the bans differed across study arms. While this study provides insight into misperceptions regarding hypothesised rationales for the bans, we did not measure differences in the perceived harmfulness of the banned versus available products. We therefore cannot be certain whether intentions to use tobacco products that remain available following the ban reflect a misperception that these products are less harmful than the banned products. This potential mechanism should be explored in future research. We also did not assess whether concurrent use of products that would remain available on the market is associated with likelihood of trying to quit in response to a ban, rather than switching to or continuing to use available tobacco products. As in other nations, many Australians who smoke use more than one type of tobacco product and there is also a high prevalence of dual use of tobacco and e-cigarettes.[28–30] The extent to which predicted behavioural responses vary according to concurrent use of products that would remain available after a ban should also be explored in future research.

## CONCLUSION

Tobacco product regulations – including bans on filter ventilation, regular nicotine content, and menthol or flavour crushball cigarettes – have significant potential to promote quit attempts among people who smoke. However, there is a need to correct and prevent misperceptions about the rationale for these regulations and the relative harm of the banned products and those that remain available, including e-cigarettes. Future research should explore the potential effectiveness of messaging strategies to induce quitting rather than product-switching in response to tobacco product regulations.

## Data availability

The dataset used in this study is not available to be shared due to conditions of ethics approval.

## Ethics approval

All study procedures were approved by the Human Research Ethics Committee of Cancer Council Victoria (HREC 2217). Informed consent was obtained from all respondents.

## Authors’ contributions

EB conceptualised and designed the study, with input from AH, SD and MW. CG conducted data analysis, with input from TN and AH. CG wrote the initial draft of the paper, with substantial review and input from AH, TN, MW, SD, and EB. All authors have checked and approved the submitted manuscript.

## Acknowledgements

The authors are grateful to all study participants and thank Ipsos for collecting these data.

